# Detection of Nucleocapsid Antibody to SARS-CoV-2 is More Sensitive than Antibody to Spike Protein in COVID-19 Patients

**DOI:** 10.1101/2020.04.20.20071423

**Authors:** Peter D. Burbelo, Francis X. Riedo, Chihiro Morishima, Stephen Rawlings, Davey Smith, Sanchita Das, Jeffrey R. Strich, Daniel S. Chertow, Richard T. Davey, Jeffrey I. Cohen

**Affiliations:** National Institute of Dental and Craniofacial Research, National Institutes of Health, Bethesda, Maryland; Medical Director Infection Control and Prevention, EvergreenHealth, Kirkland, Washington; Department of Laboratory Medicine, University of Washington, Seattle, Washington; Division of Infectious Diseases and Global Public Health, San Diego Center for AIDS Research (CFAR), University of California San Diego, San Diego, California; Department of Laboratory Medicine, Clinical Center, National Institutes of Health, Bethesda, Maryland; Critical Care Medicine Department, Clinical Center, National Institutes of Health, Bethesda, Maryland; Laboratory of Immunoregulation, National Institute of Allergy and Infectious Diseases, National Institutes of Health, Bethesda, Maryland; Laboratory of Infectious Diseases, National Institute of Allergy and Infectious Diseases, National Institutes of Health, Bethesda, Maryland

**Keywords:** COVID-19, SARS-CoV-2, coronavirus, serology

## Abstract

**Background:** SARS-CoV-2, the cause of coronavirus disease 2019 (COVID-19), is associated with respiratory-related morbidity and mortality. Assays to detect virus-specific antibodies are important to understand the prevalence of infection and the course of the immune response.

**Methodology:** Quantitative measurements of plasma or serum antibodies by luciferase immunoprecipitation assay systems (LIPS) to the nucleocapsid and spike proteins were analyzed in 100 cross-sectional or longitudinal samples from SARS-CoV-2-infected patients. A subset of samples was tested with and without heat inactivation.

**Results:** Fifteen or more days after symptom onset, antibodies against SARS-CoV-2 nucleocapsid protein showed 100% sensitivity and 100% specificity, while antibodies to spike protein were detected with 91% sensitivity and 100% specificity. Neither antibody levels nor the rate of seropositivity were significantly reduced by heat inactivation of samples. Analysis of daily samples from six patients with COVID-19 showed anti-nucleocapsid and spike antibodies appearing between day 8 to day 14 after initial symptoms. Immunocompromised patients generally had a delayed antibody response to SARS-CoV-2 compared to immunocompetent patients.

**Conclusions:** Antibody to the nucleocapsid protein of SARS-CoV-2 is more sensitive than spike protein antibody for detecting early infection. Analyzing heat-inactivated samples by LIPS is a safe and sensitive method for detecting SARS-CoV-2 antibodies.

---

Infections with severe acute respiratory syndrome coronavirus 2 (SARS-CoV-2) causing coronavirus disease 2019 (COVID-19), were first reported in China [1-4]. The major clinical feature of COVID-19 SARS-CoV-2 is virus-associated pneumonitis [5-7]. Unlike other highly pathogenic coronaviruses such as SARS/SARS-CoV-1 and Middle East respiratory syndrome coronavirus (MERS-CoV) [8], SARS-CoV-2 spreads more rapidly and reached six of the seven continents, including North America [9], within three months of the initial outbreak. Nucleic acid-based testing of oropharyngeal or nasopharyngeal swabs and saliva is useful for diagnosing acute infection. SARS-CoV-2 virus RNA can often be detected in upper respiratory secretions at the time of the first appearance of symptoms, peaks during the first week, and later declines with time [10, 11]. RNA from SARS-CoV-2, like the related SARS-CoV-1 [12], can also be detected in blood [11, 13], and high levels of circulating viral RNA are associated with more severe disease [13].

Assessment of the antibody response to SARS-CoV-2 should complement the RNA-based tests and improve our understanding of the disease course, contribute to epidemiological studies and inform vaccine development. Antibodies to the nucleocapsid protein are the most sensitive target for serologic diagnosis of infection with SARS-CoV-1 [14, 15]. Antibodies against the spike protein of SARS-CoV-1, the target of neutralizing antibody and vaccine development, emerge at a later time than those against the nucleocapsid protein. Recently, several groups have reported serological diagnostic tests using the nucleocapsid and/or spike protein from SARS-CoV-2 by ELISA [11, 16, 17], immunofluorescence [18] and even a lateral flow test [19]. One study that used ELISA to measure only antibodies to the nucleocapsid protein found that patients become seropositive 10-18 days after the onset of symptoms [16]. A commercial ELISA using the spike protein demonstrated that IgG antibodies were detectable at a median of 14 days after onset of symptoms [17]. To et al. examined antibodies against both the spike and nucleocapsid by ELISA in a small number of samples and found that IgG antibodies against the nucleocapsid protein were generally detectable at about the same time as antibodies to the spike protein [11]. Despite these findings, further studies are needed to better understand antibody dynamics in persons infected with SARS-CoV-2 to determine the most sensitive and specific antibody assays, and to use these antibody-based tests to determine seroprevalence in different populations. In addition, it is currently unknown whether the viral RNA that has been detected in the blood [11, 13] indicates the presence of infectious virus, but this has the potential to be a safety hazard for health care workers, clinical laboratory technicians and researchers analyzing serology in persons infected with SARS-CoV-2. Thus, a sensitive and specific antibody assay using heat treated plasma or serum may enhance safety when working with these fluids.

We and others have employed a liquid phase immunoassay technology termed Luciferase Immunoprecipitation Systems (LIPS) to measure antibodies against many different viruses, to stratify infected patients based on the level of their antibodies, and for virus discovery [20]. LIPS has shown promise for detecting antibodies against coronaviruses including the nucleocapsid of MERS-CoV [21] and the spike protein of swine acute diarrhea syndrome coronavirus (SADS-CoV) [22]. Unlike ELISA, which is solid phase, LIPS is performed in solution, thus maintaining the native antigen conformation. The antigen is produced in mammalian cells and often retains post-translational modifications of the antigen, unlike bacterial recombinant proteins or peptide based ELISAs. LIPS assays typically have a dynamic range up to 6 log_10_ for some antigens and require < 5 ul of plasma or sera for testing. Here recombinant nucleocapsid and spike protein from SARS-CoV-2 as antigens in LIPS assays were used to measure antibodies in patients with COVID-19 from four geographically disparate locations across the United States. The LIPS assay showed high sensitivity and specificity for detecting SARS-CoV-2 antibodies and demonstrated that nucleocapsid antibodies emerge before spike antibodies. Moreover, as there are potential safety issues related to the presence of SARS-CoV-2 RNA in blood, we show that heat inactivation of plasma at 56°C for 30 min does not significantly reduce the sensitivity of the LIPS assay and thus allows testing to be performed more safely.

## METHODS

### Characteristics of the patients with COVID-19

This retrospective study analyzed both cross-sectional and longitudinal blood samples collected from patients with COVID-19 or controls from four clinical sites. Anonymized plasma or serum from patients from University of California, San Diego (UCSD, n=3), University of Washington, Seattle (UW, n=17), EvergreenHealth, Kirkland, Washington (EH, n=23) (**Table 1**) were obtained under an IRB exemption. Plasma from patients at the NIH Clinical Center, NIH (n=6) were obtained under a protocol approved by the IRB of the NIH Intramural Research Program; all patients signed consent. Additional anonymized blood bank donor controls (n=32) collected at the NIH Clinical Center prior to 2018 were used as uninfected controls for serological testing. The time interval between the initial symptoms and obtaining plasma/serum samples from PCR+ confirmed cases was variable and ranged from 2 to 50 days. SARS-CoV2 infection was confirmed in each case by reverse transcriptase PCR detection of viral RNA from nasal and/or throat swabs performed at clinical laboratories associated with each location. Thirteen patients from the UW, 13 of 23 subjects from the outbreak at EH (including the nursing home and family members of health workers), 3 patients from the UCSD (two samples from each), and 6 patients from the NIH Clinical Center, Bethesda, MD were confirmed positive for SARS-CoV-2 RNA. In the case of the NIH samples, serial daily blood drawn samples (n=68) were available covering 0-20 days from symptom onset.

**Table 1.** Subject Characteristics of COVID-19 Cohort.

|  | N | Gender<br>(M:F) | Age<br>years | With one or<br>More Risk<br>Factors* | SARS-CoV-2<br>PCR Positive | Time from<br>Symptoms to<br>First Blood Draw | Ventilator | Mortality |
| --- | --- | --- | --- | --- | --- | --- | --- | --- |
|  |  | ratio | (range) | no. (%) | no. (%) | Average (range) | no. (%) | no. (%) |
| Blood Donors | 32 | ND | ND | ND | ND | ND | ND | ND |
| Suspected Cases<sup>#</sup> | 10 | 4:6 | 32(7-49) | 0 (0)<sup>‡</sup> | 0 (0)<sup>§</sup> | 47.1 Days (26-79)<sup>‡</sup> | 0 (0) | 0 (0) |
| Univ. Calif., San Diego | 3 | 2:1 | 73 (59-84) | 2 (66) | 3 (100) | 7.8 Days (5-14) | 3 (100) | 1 (33) |
| Univ. of Washington | 13 | 10:3 | 66 (43-95) | 13 (100) | 13 (100) | 13.2 Days (4-24) | 4 (31) | 5 (38) |
| EvergreenHealth | 13 | 3:10 | 59 (19-88) | 6 (46) | 13 (100) | 18 Days (2-50)<sup>‡</sup> | 3 (23) | 3 (23) |
| NIH Clinical Center | 6 | 5:1 | 45 (22-67) | 5 (83) | 6 (100) | 5.5 Days (0-11) | 3 (50) | 1 (17) |

\*Risk factors including heart disease, lung disease, diabetes, obesity, and/or immunocompromise
Abbreviation: ND, not determined
<sup>#</sup>EvergreenHealth
<sup>§</sup>2 PCR negative and 8 not determined
<sup>‡</sup>Unknown for 1 subject

### Storage and Heat Inactivation

Plasma/serum samples were collected and stored frozen at -80° C, except for the heat-inactivated samples from the NIH that were not previously frozen. In light of previous studies that showed a marked loss in infectivity of SARS-CoV-1 [23] and MERS [24] coronaviruses with heating, we adopted a precautionary safety protocol performed before analysis. An aliquot of plasma/serum from each patient sample was first incubated at 56° C for 30 min and then used for testing as described below.

### Luciferase Immunoprecipitation Systems (LIPS) for Measurement of SARS-CoV-2 Antibodies

LIPS assays, in which viral proteins fused to light-emitting luciferase are immunoprecipitated, were essentially performed as described [25]. A plasmid expressing the nucleocapsid of SARS-CoV-2 (amino acids 1-417 of GenBank MN908947) was generated as a synthetic DNA (Twist Biosciences) and cloned into the pREN2 eukaryotic expression vector as C-terminal *Renilla* luciferase fusion protein. A plasmid expressing the spike protein of SARS-CoV-2 (amino acids 1-538 of GenBank MN908947) was generated by PCR from a plasmid containing a prefusion form of the spike protein (2019-nCoV-2_S-2P [26]) and produced as a N-terminal fusion protein in the pGAUS3 vector for expression as a *Gaussia* luciferase fusion protein. The resulting plasmid was termed pGAUS3-Spike. A second spike construct, pGAUS3-Spike-Δ2 (amino acids 1-513) was also constructed in the pGAUS3 vector in the same way. Preliminary tests comparing antibody detection using pGAUS3-Spike-Δ2 and pGAUS3-Spike showed similar results and the former construct was not used further.

Nucleocapsid and spike protein-light emitting plasmid constructs were transfected into Cos1 cells with Fugene-6 and lysates were harvested 48 hours later to obtain crude cell extracts. For testing, heat-inactivated serum or plasma samples were diluted 1:10 in assay buffer A (20 mM Tris, pH 7.5, 150 mM NaCl, 5 mM MgCl_2_, 1% Triton X-100) and 10 μl of the diluted sample were then tested in a 96-well microtiter plate as described [25]. After incubation at room temperature for one hour, the mixture was transferred to a microtiter filter plate containing protein A/G beads and incubated for one hour. The antibody-antigen-bead complexes were then washed eight times with buffer A and twice with PBS on a microtiter filter plate to remove unbound antigens. After the final wash coelenterazine substrate (Promega) was added to detect *Renilla* luciferase and *Gaussia* reporter activity and light units (LU) were measured in a Berthold LB 960 Centro microplate luminometer (Berthold Technologies, Bad Wildbad).

Antibody levels were reported as the geometric mean level (GML) with 95% confidence interval (CI). Cut-off limits for determining positive antibodies in the SARS-CoV-2-infected samples were based on the mean plus three standard deviations of the serum values derived from the 32 uninfected blood donor controls or by receiver operator characteristics (ROC) analysis. For some of the data percentages for categorical variables, mean and range, geometric mean plus 95% CI were used to describe the data. Wilcoxon signed rank were used for statistical analysis.

## RESULTS

### Characteristics of the patients with COVID-19

Patients with COVID-19 were located in four geographically distinct locations across the United States and included 35 SARS-CoV-2 cases confirmed by PCR, 10 subjects with COVID-19-like symptoms or household contacts of persons with COVID-19 (not tested by PCR), and 32 blood donors who donated samples before 2018 used as controls (Table 1). The majority of the SARS-CoV-2 PCR-confirmed cases were male (87%) and the median age was 44 years (range, 32-50 years). A subset of the SARS-CoV-2 PCR-confirmed cases had one or more risk factors including heart disease, lung disease, diabetes, and/or they were immunocompromised. Two different plasma samples, drawn 2-3 days apart, were available for each of the three COVID-19 cases from the UCSD and multiple daily samples were available from the NIH patients with COVID-19. Combining the cross-sectional and longitudinal studies resulted in 100 samples from PCR+ patients.

### Detection of Antibodies to the Nucleocapsid Protein of SARS-CoV-2 is More Sensitive than Antibodies to the Spike Protein in COVID-19 Patients

LIPS assays for detecting antibodies were developed using SARS-CoV-2 nucleocapsid and spike antigens produced in mammalian cells. Pilot experiments using nucleocapsid-*Renilla* luciferase and spike protein-*Gaussia* luciferase fusion proteins were conducted with serum or plasma from blood donor controls collected prior to 2018. Results showed a low background with little or no antibody immunoreactivity against the spike protein, but there was a higher background immunoreactivity against the nucleocapsid (**data not shown**). Based on the need to develop a highly specific SARS-CoV-2 LIPS serological test without potential false positives, stringent cut-off values from the blood donor controls were assigned based on statistical methods and/or ROC. From this analysis, cut-off values for the nucleocapsid and spike proteins were derived from the mean plus four standard deviations (125,000 LU) and the mean plus three standard deviations (45,000 LU) of the blood donor controls, respectively.

Using these cut-off values, plasma or serum samples from the COVID-19 cohort were evaluated by LIPS assay for antibodies against the nucleocapsid or spike protein. For safety reasons, all samples used for this analysis were heated at 56°C for 30 min to reduce the likelihood of having infectious virus in the samples. Coded plasma or serum samples from suspected COVID-19 cases from EH were then tested as well as noncoded pre-2018 blood donors, and SARS-CoV-2 PCR-positive cases from the UCSD, UW, EH, and the NIH Clinical Center (NIH). A wide dynamic range of antibody levels against the nucleocapsid and spike protein were observed differing by up to 100-fold between samples (**Figure 1**). In order to compare the sensitivity of the nucleocapsid and spike LIPS assays, a minimum interval of >14 days between onset of symptoms and time of blood collection was used to determine the number of seropositive serum or plasma samples in the SARS-CoV-2 PCR-positive group. Among the PCR+ patient samples collected >14 days after onset of symptoms (**Figure 1, black dots**), seropositive nucleocapsid antibodies were detected in 4/4 patients from UW, 7/7 from EH, and 32/32 serial samples from the six NIH patients, yielding both a sensitivity and specificity of 100%. A similar analysis of the spike antibody in samples collected >14 days after onset of symptoms showed a slightly lower sensitivity of 91% (32/35) with 100% specificity, where 4/4 patients from UW, 6/7 from EH, and 22/24 from NIH were seropositive.

**Figure 1.**
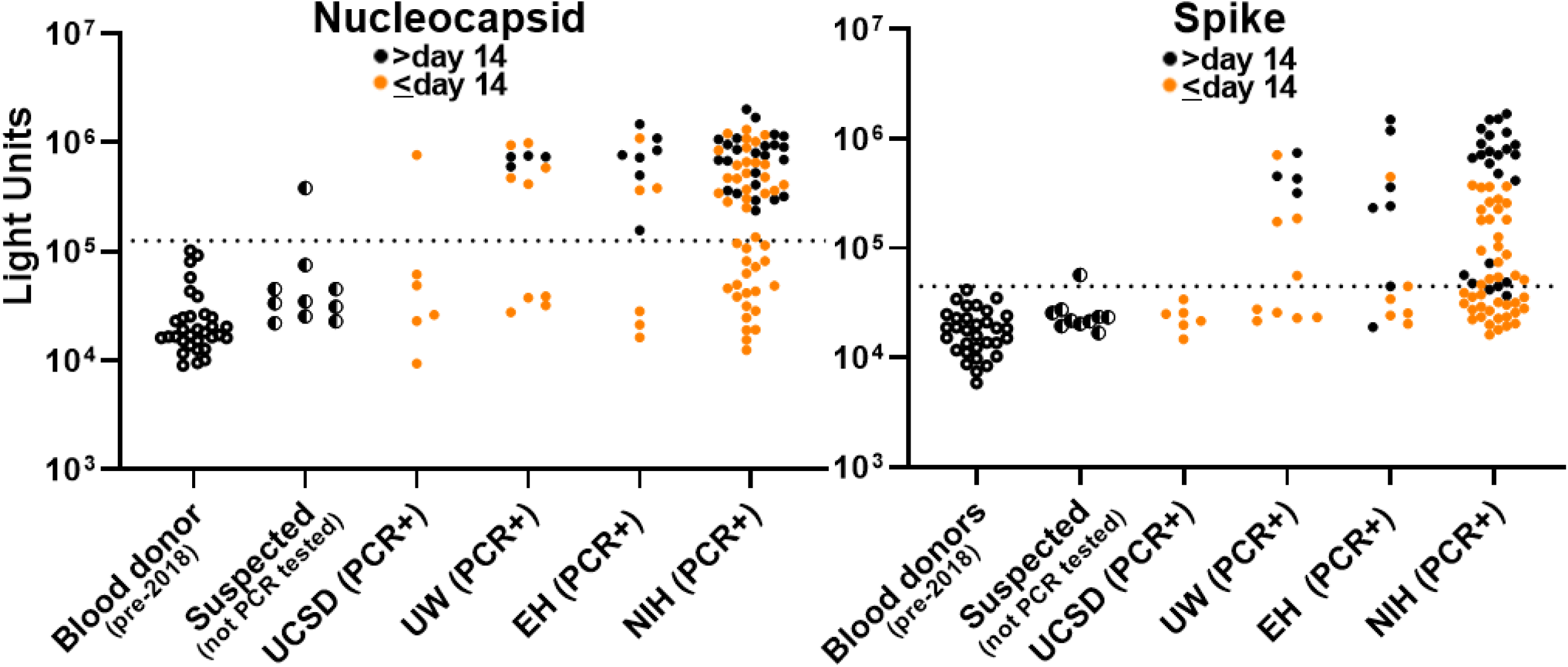
Detection of antibodies against SARS-CoV-2 nucleocapsid and spike protein in patients with COVID-19. Antibody levels against SARS-CoV-2 nucleocapsid and spike protein were determined in 32 pre-2018 blood donors, 10 suspected COVID-19 cases (not PCR confirmed) from EvergreenHealth, Kirkland, WA (EH), three PCR+ COVID-19 patients from UCSD, 13 PCR+ COVID-19 patients from the University of Washington (UW), 13 PCR+ COVID-19 patients from EH, and 6 COVID-19 patients from the NIH Clinical Center (NIH). Each symbol represents a sample from an individual patient or different time points from an individual patient. Antibody levels are plotted in light units (LU) on a log_10_ scale. Black circles represent plasma or serum samples obtained after 15 or more days after symptom onset and orange circles are from plasma or serum samples obtained 14 or less days after symptom onset. The dashed lines represent the cutoff level for determining positive antibody titers as described in the Methods.

Evaluation of samples collected at ≤14 days after onset of symptoms showed reduced sensitivity, but specificity was maintained. The sensitivity for antibody to the nucleocapsid protein at this time point was 51% (33/65) with antibodies detected in 1/6 samples from UCSD, 5/9 from UW, 3/6 from EH and 24/44 from NIH (**Figure 1, orange dots**). Analysis of spike antibodies of samples collected at ≤14 days after onset of symptoms showed a sensitivity of 43% (28/65) with antibody detected in 0/6 samples from UCSD, 4/9 samples from UW, 2/6 from EH and 22/44 from NIH. Taken together, our findings indicate that detection of antibodies against the nucleocapsid protein is more sensitive than detection of antibodies against the spike protein, and that nucleocapsid antibodies generally appear earlier than spike antibodies.

In addition to the SARS-CoV-2 PCR-confirmed patients, suspected cases of COVID-19 from EH were also analyzed for seropositivity. Nine of the ten suspected cases without viral PCR confirmation, that showed symptoms compatible with COVID-19 collected between January and February 2020, were seronegative for both nucleocapsid and spike antibodies (**Figure 1**). Interestingly, one case from March 2020 from a person who was a household contact with a SARS-CoV-2 PCR+ patient, was seropositive for both nucleocapsid and spike antibodies.

Since there is interest in using serological assays to assess current and historical infections, we evaluated the robustness of the LIPS assay for detecting SARS-CoV-2 antibodies by analyzing the level of antibodies in all PCR-confirmed samples collected more than 14 days after symptom onset. The geometric mean level (GML) of nucleocapsid antibody levels in the 35 seropositive samples was 694,600 (95% CI, 570,000-844,600 LU), which was approximately 32 times higher than the GML of the blood donor controls of 21,356 LU (95% CI, 17,032-26,752). Antibodies against spike protein showed a similar discriminatory potential for the seropositive samples with GML of 346,800 LU (95% CI, 218,800-550,000 LU), which was approximately 21 times higher than the blood donor controls with 16,843 LU (95% CI, 14,172-20,007 LU). These findings indicate that the LIPS assays for antibodies to nucleocapsid and spike protein are robust and should be useful to evaluate the prevalence of infection with. SARS-CoV-2.

### Time course of the appearance of serum antibodies against SARS-CoV-2 differs in immunocompetent and immunocompromised patients

To understand the timing and trajectory of SARS-CoV-2 antibodies against nucleocapsid and spike proteins, serial daily blood samples from the six NIH patients with COVID-19 were studied. In all six subjects, SARS-CoV-2 antibody levels rose with time in both the three immunocompetent (**Figure 2A**, NIH patients 1-3) and three immunocompromised patients (**Figure 2B**, NIH patients 4-6). These latter three patients had chronic lymphocytic leukemia, metastatic chordoma, or had received a hematopoietic stem-cell transplant. All three immunocompetent COVID-19 patients showed a rapid rise in antibody to nucleocapsid and began within 10 days of symptom onset in 2 patients (no samples were available before day 11 for the third patient, **Figure 2A)**. Antibodies against the spike protein in these three immunocompetent patients generally tracked with the nucleocapsid antibodies, but in one case seropositivity appeared 2 days later than nucleocapsid antibody. The third patient, NIH-3, with a history of hypertension and heart disease died of cardiovascular shock and hypoxemia 13 days after onset of symptoms.

**Figure 2.**
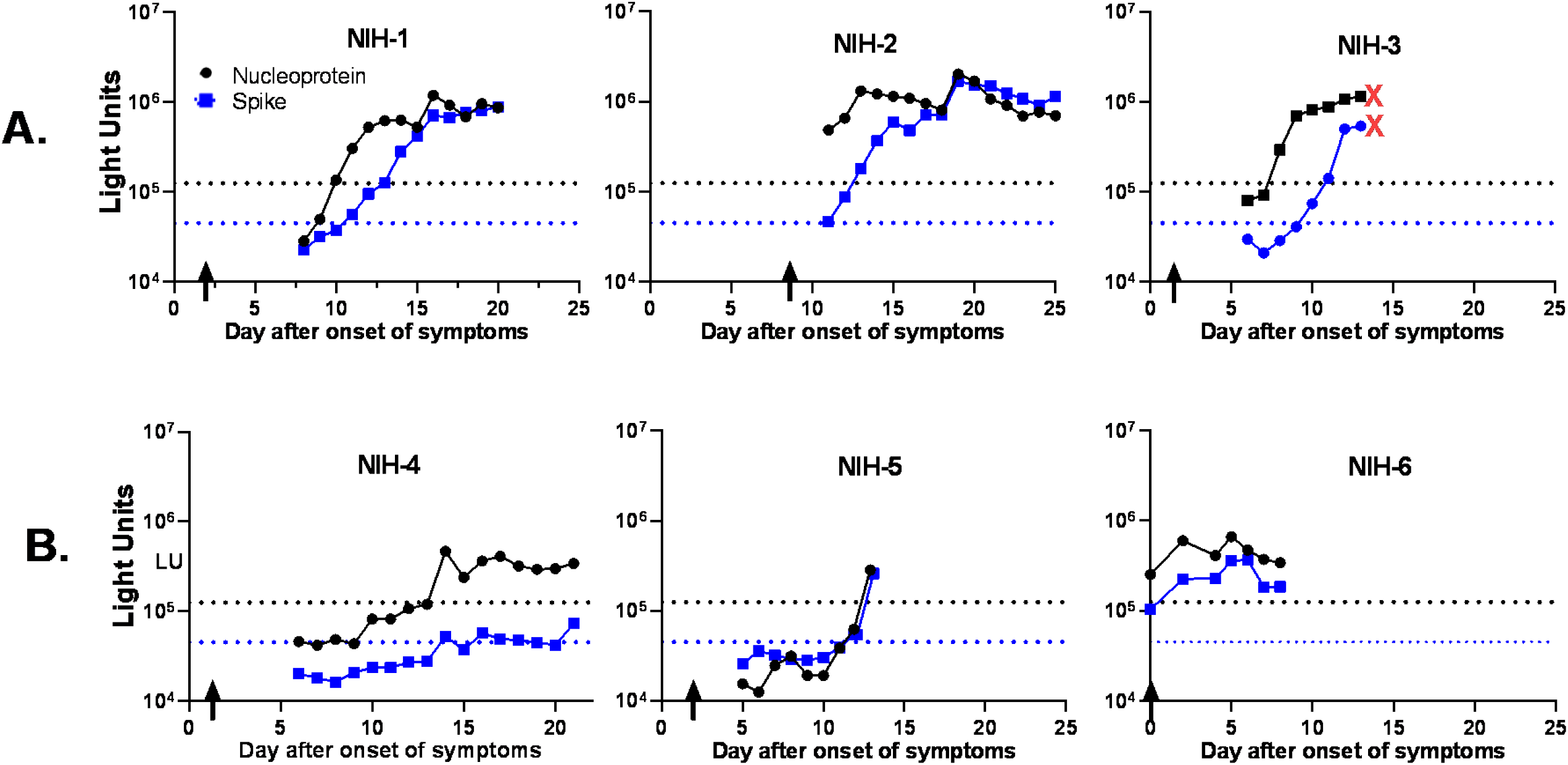
Longitudinal profile of antibodies against nucleocapsid and spike protein in immunocompetent and immunocompromised COVID-19 patients from NIH. Antibody levels were determined in daily blood draws from six COVID-19 patients. Three of the COVID-19 patients were immunocompetent (**Panel A, NIH-1-3**) and three (**Panel B, NIH-4-6)** were immunocompromised. The levels of antibody to the nucleocapsid (black line) and spike protein (blue line) over time are shown and were plotted on the y-axis using a log_10_ scale. Time zero represents the first day symptoms appeared, and the vertical arrows are the time of diagnosis by PCR. The cut-off values for determining seropositivity is shown by the dotted lines. The red X’s indicates the day after onset of symptoms that patient NIH-3 died.

Antibody profiles in the three immunocompromised NIH patients showed more blunted responses against the SARS-CoV-2 antigens (**Figure 2B**). Patient NIH-4 became seropositive for both nucleocapsid and spike antibodies on day 14 and these antibodies then plateaued at these low levels for the next seven days. Similarly, patient NIH-5 did not become seropositive until day 13 for spike antibody and day 14 for nucleocapsid antibody. Patient NIH-6 was both PCR+ for SARS-CoV-2 and seropositive on the day of symptoms, suggesting that he had an asymptomatic infection for several days before diagnosis. Despite the blunted antibody response, none of the immunocompromised patients died. Overall, the results with this small group of patients suggests that immunocompromised patients generally have a more attenuated and/or delayed antibody response to SARS-CoV-2 than immunocompetent patients.

### Heat inactivation of plasma minimally impacts detection of antibody to SARS-CoV-2 in the LIPS assay

While heating plasma to 56° C for 30 min has been shown to reduce the titer of human coronaviruses, heating might reduce or eliminate IgM and IgG responses [27]. Therefore, we performed LIPS assays on a subset of the patients with COVID-19 from the known or suspected cases (N=38) with and without heat inactivation, to evaluate its impact on nucleocapsid antibody levels and seropositivity status. Evaluation of antibody responses in heated versus unheated plasma samples showed that antibody levels were mostly unchanged (**Figure 3**). In a single sample from one patient with COVID-19, antibody to SARS-CoV-2 was not detected after heat-inactivation. Of note, this sample came from an NIH patient with COVID-19 who was antibody positive at day 7 using non-heated plasma and became seropositive using heat-inactivated plasma from day 8. Statistical analysis showed no significant difference in antibody levels between plasma that was heated or unheated (Wilcoxon Signed rank test) and the values were highly correlative (Rs=0.913; P<0.0001). These findings indicate that the heat-inactivation process is diagnostically suitable for testing of SARS-CoV-2 antibodies by LIPS.

**Figure 3.**
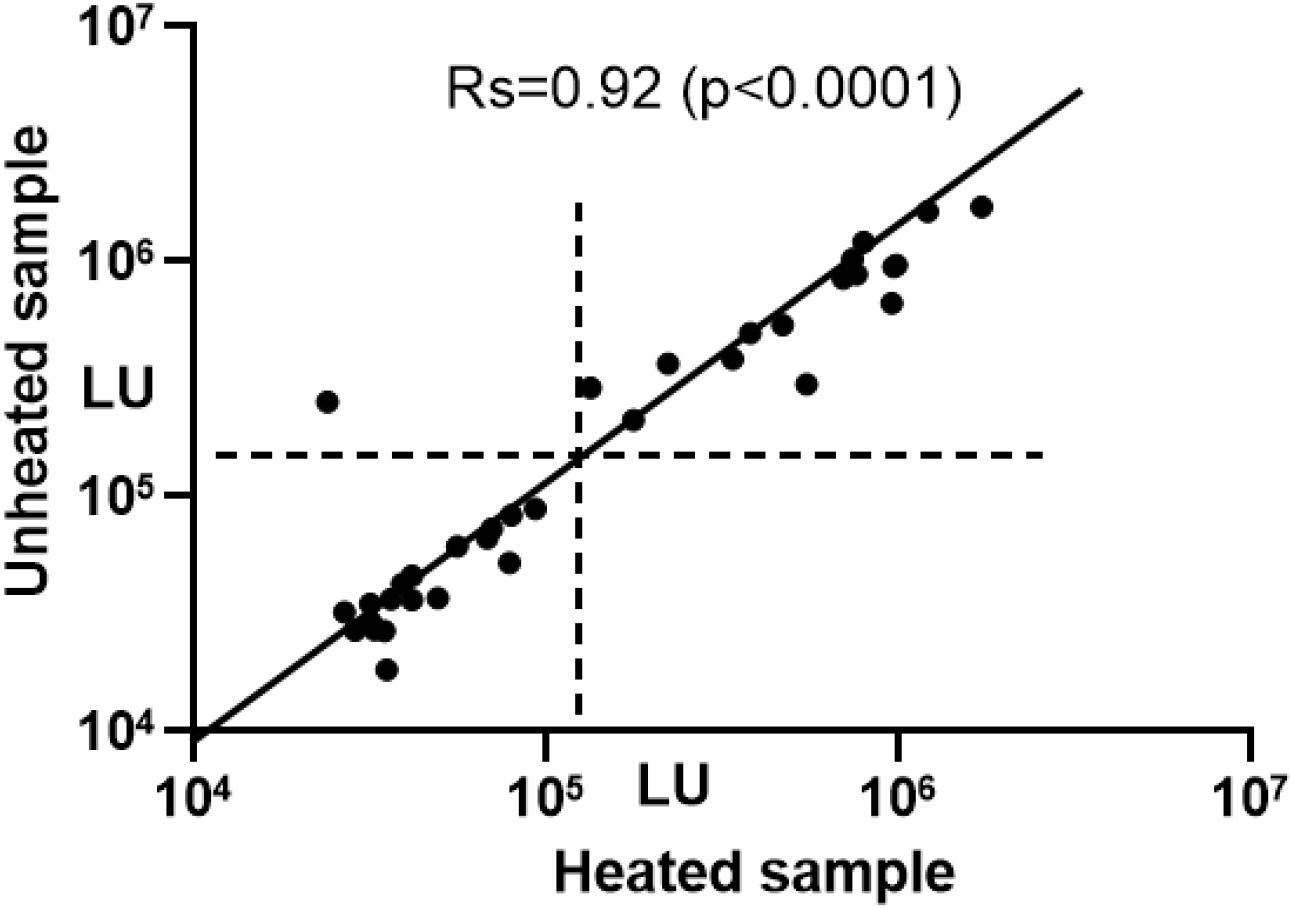
Heat inactivation of plasma or serum samples has no significant impact on detection of nucleocapsid antibodies. A subset (n=38) of plasma samples from patients with COVID-19 including samples from PCR-positive patients from very early infection (less than 8 days) and at later times after initial infection were analyzed. Levels of antibody to the nucleocapsid protein were determined by LIPS for aliquots of paired samples from unheated plasma or serum and from heated plasma or serum. Antibody levels were plotted, and the horizontal and vertical dotted lines represent the cutoff values for seropositivity. The diagonal line is a theoretical value if the antibody levels were identical for heated and unheated samples. The antibody values strongly correlated for heat treated and not heat-treated samples as shown by the Spearman rank correlation (*Rs*) of 0.92 (*P <*0.0001) and only one sample showed a significant decrease with heating.

## DISCUSSION

We used a fluid-phase LIPS assay to investigate antibodies to the SARS-CoV-2 nucleocapsid and spike protein in COVID-19 patients after infection. The LIPS assay demonstrated high sensitivity and a wider dynamic range for antibody detection compared to other published assays [11, 16-19]. An analysis of longitudinal plasma samples showed that antibodies against the nucleocapsid and spike proteins appeared about the same time between day 8 and day14 after the onset of symptoms. Only one study to date has examined antibodies separately against the nucleocapsid protein and spike protein [11] and our findings are in general agreement. COVID-19 patient plasma samples obtained ≥ 14 days after symptom onset showed that the LIPS assay for antibodies against the nucleocapsid and spike protein had 100% and 94% sensitivity, respectively, with 100% specificity for both antibodies. Additional studies using this high-throughput, highly quantitative LIPS assay may also help determine whether the relative levels of antibodies observed in convalescent COVID-19 patients or uninfected vaccinated persons correlate with prevention of reinfection or primary infection, respectively. Quantitative antibody profiles will be useful in determining antibody decay over time. It is known that for some viral infections there is long-lasting antibody responses and protection from infection, but for others antibody levels wane at faster rates [28]. Following humoral response profiles of natural infection from convalescent COVID-19 cases over time should provide important insights into the half-life of these antibodies.

Using the quantitative LIPS assay, our studies with serial patient samples from the NIH cohort showed the temporal relationship between antibody dynamics with onset of symptoms and PCR positivity for SARS-CoV-2. Cut-off values for a positive result was based on pre-2018 blood donors and may underestimate the number of seropositive persons because some individual patients showed low antibody values initially that gradually rose before exceeding the cut-off value. Nevertheless, all three of the immunocompetent COVID-19 patients showed rapid seroconversion within 10 days of onset of symptoms for antibody to the nucleocapsid protein and robust, but slightly delayed for antibody to the spike proteins. In contrast, the immunocompromised NIH patients exhibited a slower rise in antibody levels with a plateau at lower levels compared to the immunocompetent patients, and two patients did not become seropositive until 14 days after onset of symptoms. Nonetheless, the immunocompromised patients had a favorable clinical outcome. The NIH patient who died (NIH-3) was not immunocompromised and had a rapid rise in antibody production reaching levels comparable to the other immunocompetent patients. In addition, one of the two EH patients who died showed the highest antibody levels in that cohort of patients. While excessive proinflammatory responses to the virus have been reported to contribute to poor outcomes [29-31], larger studies of COVID-19 patients are required to determine whether antibody levels directly correlate with disease severity.

Prior studies have shown high levels of SARS-CoV-2 RNA in blood from patients with COVID-19 [1, 4]. At present, it is not certain whether infectious virus might be circulating in the blood early during infection. Accordingly, we heated plasma or serum to 56° C for 30 min to reduce the titer of SARS-CoV-2 before performing the LIPS assays, since prior studies have shown a marked loss in infectivity of SARS-CoV-1 [23, 32] and MERS [24] coronaviruses with heat treatment. While impaired detection of viral IgM and IgG antibody responses to viruses after heating samples to 56°C has been reported [27], and several abstracts report similar findings with SARS-CoV-2 samples, our direct comparison of untreated and treated samples found high concordance of the antibody values revealing the suitability of heat inactivation. This inactivation protocol may be useful to enhance safety when studying highly infectious saliva from COVID-19 patients [11] for IgG and IgA antibodies. Further modification of LIPS assays for detection of SARS-CoV-2 antibodies, including the use of different protein fragments, full-length spike protein, and/or different luciferase reporters, may further improve assay performance. Nonetheless, our current assay provides highly quantitative results with a high degree of sensitivity and specificity and should be useful for larger seroepidemiologic studies.

## Data Availability

Full data will be available when the manuscript is published

## Acknowledgements

We thank Kizzmekia S. Corbett, Barney Graham, and Jason S. McClellan for the gift of plasmid 2019-nCoV-2_S-2P, Jocelyn Voell for assistance with sample collection, Marie K. Hensley, Jemilin E. Mendoza and Kristine A. Vander Hart for clinical data management, and Jason Van Winkle, MD and Diego Lopez de Castilla, MD for clinical support

## References

1. Chan JF, Yuan S, Kok KH et al. A familial cluster of pneumonia associated with the 2019 novel coronavirus indicating person-to-person transmission: a study of a family cluster. Lancet 2020; 395:514–23.

2. Wu F, Zhao S, Yu B et al. A new coronavirus associated with human respiratory disease in China. Nature 2020; 579:265–69.

3. Zhou P, Yang XL, Wang XG et al. A pneumonia outbreak associated with a new coronavirus of probable bat origin. Nature 2020; 579:270–73.

4. Chen L, Liu W, Zhang Q et al. RNA based mNGS approach identifies a novel human coronavirus from two individual pneumonia cases in 2019 Wuhan outbreak. Emerg Microbes Infect 2020; 9:313–19.

5. Chen N, Zhou M, Dong X et al. Epidemiological and clinical characteristics of 99 cases of 2019 novel coronavirus pneumonia in Wuhan, China: a descriptive study. Lancet 2020; 395:507–13.

6. Huang C, Wang Y, Li X et al. Clinical features of patients infected with 2019 novel coronavirus in Wuhan, China. Lancet 2020; 395:497–506.

7. Zhu N, Zhang D, Wang W et al. A Novel Coronavirus from Patients with Pneumonia in China, 2019. N Engl J Med 2020; 382:727–33.

8. Cui J, Li F, Shi ZL. Origin and evolution of pathogenic coronaviruses. Nat Rev Microbiol 2019; 17:181–92.

9. Holshue ML, DeBolt C, Lindquist S et al. First Case of 2019 Novel Coronavirus in the United States. N Engl J Med 2020; 382:929–36.

10. Zou L, Ruan F, Huang M et al. SARS-CoV-2 Viral Load in Upper Respiratory Specimens of Infected Patients. N Engl J Med 2020; 382:1177–79.

11. To KK, Tsang OT, Leung WS et al. Temporal profiles of viral load in posterior oropharyngeal saliva samples and serum antibody responses during infection by SARS-CoV-2: an observational cohort study. Lancet Infect Dis 2020.

12. Chang L, Yan Y, Wang L. Coronavirus Disease 2019: Coronaviruses and Blood Safety. Transfus Med Rev 2020.

13. Chen W, Lan Y, Yuan X et al. Detectable 2019-nCoV viral RNA in blood is a strong indicator for the further clinical severity. Emerg Microbes Infect 2020; 9:469–73.

14. Leung DT, Tam FC, Ma CH et al. Antibody response of patients with severe acute respiratory syndrome (SARS) targets the viral nucleocapsid. J Infect Dis 2004; 190:379–86.

15. Zhu H, Hu S, Jona G et al. Severe acute respiratory syndrome diagnostics using a coronavirus protein microarray. Proc Natl Acad Sci U S A 2006; 103:4011–6.

16. Guo L, Ren L, Yang S et al. Profiling Early Humoral Response to Diagnose Novel Coronavirus Disease (COVID-19). Clin Infect Dis 2020.

17. Zhao J, Yuan Q, Wang H et al. Antibody responses to SARS-CoV-2 in patients of novel coronavirus disease 2019. Clin Infect Dis 2020.

18. Wolfel R, Corman VM, Guggemos W et al. Virological assessment of hospitalized patients with COVID-2019. Nature 2020.

19. Li Z, Yi Y, Luo X et al. Development and Clinical Application of A Rapid IgM-IgG Combined Antibody Test for SARS-CoV-2 Infection Diagnosis. J Med Virol 2020.

20. Burbelo PD, Lebovitz EE, Notkins AL. Luciferase immunoprecipitation systems for measuring antibodies in autoimmune and infectious diseases. Transl Res 2015; 165:325–35.

21. Alagaili AN, Briese T, Mishra N et al. Middle East respiratory syndrome coronavirus infection in dromedary camels in Saudi Arabia. mBio 2014; 5:e00884–14.

22. Zhou P, Fan H, Lan T et al. Fatal swine acute diarrhoea syndrome caused by an HKU2-related coronavirus of bat origin. Nature 2018; 556:255–58.

23. Yunoki M, Urayama T, Yamamoto I, Abe S, Ikuta K. Heat sensitivity of a SARS-associated coronavirus introduced into plasma products. Vox Sang 2004; 87:302–3.

24. Leclercq I, Batejat C, Burguiere AM, Manuguerra JC. Heat inactivation of the Middle East respiratory syndrome coronavirus. Influenza Other Respir Viruses 2014; 8:585–6.

25. Burbelo PD, Ching KH, Klimavicz CM, Iadarola MJ. Antibody profiling by Luciferase Immunoprecipitation Systems (LIPS). J Vis Exp 2009; Oct 7;(32).

26. Wrapp D, Wang N, Corbett KS et al. Cryo-EM structure of the 2019-nCoV spike in the prefusion conformation. Science 2020; 367:1260–63.

27. Al-Muzairai IA, Mansour M, Almajed L et al. Heat inactivation can differentiate between IgG and IgM antibodies in the pretransplant cross match. Transplant Proc 2008; 40:2198–9.

28. Amanna IJ, Carlson NE, Slifka MK. Duration of humoral immunity to common viral and vaccine antigens. N Engl J Med 2007; 357:1903–15.

29. Chen T, Wu D, Chen H et al. Clinical characteristics of 113 deceased patients with coronavirus disease 2019: retrospective study. BMJ 2020; 368:m1091.

30. Chen G, Wu D, Guo W et al. Clinical and immunologic features in severe and moderate Coronavirus Disease 2019. J Clin Invest 2020.

31. Guo T, Fan Y, Chen M et al. Cardiovascular Implications of Fatal Outcomes of Patients With Coronavirus Disease 2019 (COVID-19). JAMA Cardiol 2020.

32. Darnell ME, Taylor DR. Evaluation of inactivation methods for severe acute respiratory syndrome coronavirus in noncellular blood products. Transfusion 2006; 46:1770–7.

